# Lack of Trust, Insufficient knowledge and Risk denial; an in-depth Understanding of Health workers Barriers to uptake of the Covid-19 vaccine at Iganga Hospital Eastern Uganda, and Mengo Hospital Kampala Uganda

**DOI:** 10.1101/2021.10.13.21264920

**Authors:** Lubega Muhamadi, Namulema Edith, Waako James, Nazarius Mbona Tumwesigye, Safinah Kisu Museene, Hellen Mukakaarisa, Stefan Swartling Peterson, Anna Mia Ekström

## Abstract

Covid 19 Vaccine hesitancy among health workers remains a major hindrance to the governments vaccine roll out plan among health workers and other target populations in Uganda.

We conducted 12 focus group discussions and 20 in-depth interviews with health workers (vaccinated and unvaccinated) to understand barriers to vaccine acceptance in their own perspective and context in central and eastern Uganda. Reported barriers to vaccine acceptance included: gross lack of trust, fear of side effects, risk denial and insufficient information about the vaccine amidst negative publicity about the vaccine from the internet and social media platforms. Others were health system inhibition factors and religious beliefs against the vaccine.

We recommend a health work context specific information, education and dissemination strategy to create awareness, information and more knowledge about the vaccine to health workers.

We also recommend a sustained government media campaign to give more information about the vaccine and also dispel the negative publicity and misinformation about the vaccine.

Dialogue with health workers at all levels of care, positive peer influence, use of religious and opinion leaders as well as government ensuring accessibly to various Covid 19 vaccines and putting vaccine posts outside hospital settings to limit exposure to Covid patients could also increase uptake of the vaccine among health workers.

## 1.0 Background

Although the government of Uganda continues to receive/procure Covid-19 vaccines for its people, vaccine hesitancy poses a major threat to the government’s rollout plan for vaccination against Covid-19 [1-3]. Uganda with a population of approximately 45 million people targeted to fully vaccinate 22 million people (49.6% of the population) in a phased manner [1, 4]. The first phase was anticipated to vaccinate 4.8 million (20%) of the target population between March and June 2021 including; health workers, security personnel, people above 50 years, below 50 years but with underlying co-morbidities, teachers and students aged 18 years and above. However, according to the presidential press release of 22^nd^ September 2021, 81% of the targeted 4.8 million people had not received their first dose, while 91% had not received their second dose of the vaccine [5]. This is against the background that the government also planned to import and administer another estimated twelve million doses of the Covid-19 vaccine by December 2021. [4, 5].

Health workers who are the most vulnerable frontline staff in the fight against Covid-19 have not fully embraced the vaccination exercise. For example, by the end September 2021, approximately 30% of the health workers had not received their first dose while 60% had not received their second dose of the Covid-19 vaccine as well [4-6]. In Mengo hospital, located at the centre of capital city Kampala, the office of the deputy director of the hospital estimates that 40%-50% of the health workers did not take up the vaccine even when it was available [7]. Similarly, in Iganga hospital eastern Uganda approximately 120 kilometers from the capital city Kampala, the vaccine uptake rate among the health workers was approximately 50% [8]. Health workers are trusted people and communities follow their example and health actions almost religiously. Therefore as the government vaccine roll out plan evolves, the health worker hesitancy towards vaccine uptake is likely to negatively impact how the general community takes up the vaccine.

Why would health workers who should lead by example, and are aware of the damage the pandemic has caused to their communities let alone seeing colleagues die of Covid-19 refuse to take up the vaccine? A few online surveys or pre vaccine era studies have shown that social influence, religious and cultural beliefs, fear of side effects, attitude towards the vaccine, susceptibility to illness among others are barriers to uptake of vaccines in general [9-12]. There is, however, paucity of information on why health workers in Uganda have not fully embraced the Covid-19 vaccine from their own context and perspective, in spite of the Covid-19 deaths among themselves, and the clients they serve. An in-depth understanding of the health workers perceptions towards this vaccine from their own perspective is therefore vital especially at a time when the government is rolling out vaccination to over 20 million people and hence this study.

## 2.0 Materials and Methods

### Study Setting and population

Between June and August 2021, we sought an in-depth understanding of the health workers perceptions with regard to the Covid-19 vaccine in Mengo and Iganga hospitals. Mengo hospital is a private not for profit entity located in the capital city Kampala. The hospital offers both general clinical and specialized services to an urban population of over three million people from the Kampala metropolitan area. The hospital employs over 800 staff consisting of technical, administrative and support staff. It is one of the designated Covid-19 treatment centres in Kampala offering both general clinical care and supportive treatment to people infected with Covid-19. It also has a high dependency unit (HDU) for clients that need high level monitoring and an intensive care unit (ICU) for critical cases. Over the first and second wave of the Covid-19 pandemic in Uganda, the hospital treated over 300 patients of different levels of severity of the infection. The hospital also serves as a Covid-19 vaccination centre [7]. Iganga hospital is located in the east of Uganda approximately 120 km east of the capital city Kampala. It is a public general referral hospital offering both general clinical and specialized services. The hospital is a referral centre for about six districts constituting a population of about three million people. Most of the patients here are predominantly rural living on subsistence farming with only approximately 7% living in urban and peri-urban environments. The hospital has approximately 200 staff consisting of technical, administrative and support staff. It is one of the designated Covid-19 treatment centres in the region offering both general clinical care and supportive treatment to people infected with Covid-19 but also has a HDU. Over the first and second wave of the Covid-19 pandemic in Uganda, the hospital treated over 200 patients of different levels of severity of the infection, fifteen of whom were health workers. The hospital also serves as a Covid-19 vaccination centre [8, 13, 14].

This qualitative study employed 12 focus group discussions (FGDs) comprising staff of different cadres and deployment. Six of the FGDs were conducted in Mengo hospital while another six were conducted in Iganga hospital. We also conducted 20 in depth interviews (IDIs) with health workers from both Mengo and Iganga hospitals 10 of whom had been vaccinated with the Covid-19 vaccine and 10 had not been vaccinated. The study aimed at exploring possible motivators/barriers for uptake of the vaccine amongst the health workers from their own perspective [15, 16]. The FGD and IDI respondents were purposively selected based on maximum variety sampling from a sampling frame covering all levels of cadres from specialist health personnel to the lowest support staff in order to elicit different viewpoints. The respondents were selected because they were considered to be more “knowledge rich” on the study subject in their own situations than anybody else [15-18]. The participants were chosen from consenting members in both hospitals. The FGDs were stratified by cadre to encourage more active participation and included those who had been vaccinated and those who had not been vaccinated to avoid potential stigmatization. Participants were invited to take part in only one FGD or IDI. Each FGD consisted of a maximum of 12 participants. While each of the participants knew their vaccination status, the participants were not told how their groups were selected or their vaccination status to avoid “playing to type”

### Data collection tools and methods

Using a topic guide, the selected participants were probed on their knowledge, attitudes and perceptions about the Covid-19 vaccine. Their views on values and norms related to risk/benefit of vaccines and public health interventions, acceptance and uptake of vaccines in general and the Covid-19 vaccine in particular, as well as prevailing common misconceptions and myths about the vaccine among others were also probed. Interviews were stopped when it was judged that saturation had been reached and no more new information could be retrieved from the respondents. All data collection was supervised and assessed by the first author LM) who is a male, indigenous public health physician and the second and second last authors (WJ, NE) both of whom are conversant with qualitative research and the health system dynamics for both Iganga and Mengo hospitals respectively. Five research assistants moderated and took notes for the study. The research assistants were conversant with qualitative data collection methods, had conducted similar research in the study setting and were all fluent in Luganda and Lusoga (the local languages). They were trained for two days on the study aim, design and tools. Role-plays were used to prepare the research assistants for their interaction with the informants [17, 19, 20]. The experiences from the role-plays were discussed and further methodological guidance was given.

### Data management and analysis

All the FGDs and the IDIs were conducted in a mixture of English, Lusoga or Luganda (respective native languages), transcribed and later all translated into English by the interviewers. The authors listened to the audio recordings to confirm the validity of the information. Data collection stopped when information relating to the topic guides revealed no new information. Data analysis was iterative including reviews and discussions at different stages of data collection and appropriate modifications were made in the tools to address emerging issues [18, 21]. The units of analysis were the transcripts from the IDIs and the FGDs. Content analysis was used to analyze the transcripts. This entailed reading and reviewing texts of the entire interview back and forth to identify meaningful units in relation to the study subject [16, 20, 21]. The meaningful units were condensed and coded by categories and themes, and discussed by (LM, WJ and EN) until consensus was reached on the appropriate codes and the themes [20].

### Ethical clearance

The study was approved by the Mengo hospital Research and Ethics Committee (REC) ref MH/REC/39/06-2021, and the Uganda National Council for Science and Technology (UNCST). We also sought the approval of the management of both Mengo and Iganga hospitals. As part of the informed consent, the participants were told about the aims of the study, the anticipated benefits and risks, their ability to participate or withdraw at any time, and assured that all information obtained would be kept confidential. The participants signed two copies of the consent form before the interview commenced, and one copy was given to the participant.

## 3.0 Results

**Table 1:** Summary characteristics of the FGD participants stratified by cadre or unit of health care.

| **FGD number** | **Number of participants** | **Age range/years** | **Sex** | **Vaccination status** |
| --- | --- | --- | --- | --- |
| 01 | 12 | 28-52 | Males and females | Vaccinated and un vaccinated |
| 02 | 12 | 26-50 | Males and females | Vaccinated and un vaccinated |
| 03 | 12 | 33-52 | Males and females | Vaccinated and un vaccinated |
| 04 | 12 | 38-58 | Males and females | Vaccinated and un vaccinated |
| 05 | 12 | 33-52 | Males and females | Vaccinated and un vaccinated |
| 06 | 12 | 38-58 | Males and females | Vaccinated and un vaccinated |
| 07 | 12 | 28-43 | Males and females | Vaccinated and un vaccinated |
| 08 | 12 | 24-40 | Males and females | Vaccinated and un vaccinated |
| 09 | 12 | 28-48 | Males and females | Vaccinated and un vaccinated |
| 10 | 12 | 20-33 | Males and females | Vaccinated and un vaccinated |
| 11 | 12 | 33-44 | Males and females | Vaccinated and un vaccinated |
| 12 | 12 | 36-65 | Males and females | Vaccinated and un vaccinated |

**A total of 144 health workers participated in the focus group discussions age range 20 to 65 years with median age of 36 years**

**Table 2:** Summary Characteristics of the IDI participants purposively identified by cadre.

| **Participants**<br>**number** | **Age range/years** | **Sex** | **Vaccination status** |
| --- | --- | --- | --- |
| 01 | 31-35 | Male | Vaccinated |
| 02 | 31-35 | Female | Vaccinated |
| 03 | 46-50 | Male | Un vaccinated |
| 04 | 36-40 | Male | Un vaccinated |
| 05 | 36-40 | Male | Un vaccinated |
| 06 | 41-45 | Female | Vaccinated |
| 07 | 36-40 | Female | Vaccinated |
| 08 | 31-35 | Male | Unvaccinated |
| 09 | 26-30 | Male | Un vaccinated |
| 10 | 26-30 | Male | Vaccinated |
| 11 | 36-40 | Female | Vaccinated |
| 12 | 31-35 | Female | Un vaccinated |
| 13 | 36-40 | Female | Un vaccinated |
| 14 | 26-30 | Female | Vaccinated |
| 15 | 26-30 | Female | Vaccinated |
| 16 | 31-35 | Female | Vaccinated |
| 17 | 46-50 | Male | Un vaccinated |
| 18 | 46-50 | Female | Un vaccinated |
| 19 | 31-35 | Female | Un vaccinated |
| 20 | 51-55 | Female | Vaccinated |

**Twenty participants were identified for the in depth interviews. Twelve were females and 8 males. Age range was 28 to 55 years with a median of 36 years.**

Our analysis of textual data generated six themes including: (1) gross lack of trust in the vaccine, (2) fear of real and perceived side effects, (3) insufficient sensitization of health workers amidst vaccine negative publicity (4) risk denial, (5) system inhibition factors and (6) religious beliefs against vaccination.

### Gross lack of trust in vaccine

Health workers expressed lack of trust in the AstraZeneca vaccine based on several unanswered questions in view of what they knew about vaccines. There concerns varied from why government was obtaining consent for the Covid-19 vaccine contrary to common practice for other vaccines, the short duration of research preceding the recommendation of the Covid-19 vaccine contrary to common practice and whether this indeed was not a vaccine trial on Ugandan themselves being used as Guinea pigs.

*“This is clearly a clinical trial and we are being duped otherwise why government would ask us to consent for it, something we have never done for other vaccines. Ugandans are being used as* guineapigs………. *so terrible” (male informant, unvaccinated)*.

*“Vaccine trials take about three years; do you really believe that a vaccine can be truly recommended for global use after testing for only a year? Something is certainly not right here” (female informant, unvaccinated)*.

The government’s failure to accept, explain or even take liability for the vaccines side effects/adverse events. As well as the public knowledge that government had accepted to use Astra Zeneca which had been branded as dangerous and rejected in other countries but dumped in Uganda further eroded trust for the vaccine among health workers.

*“*..*if the government is confident about the vaccine, why aren’t they explaining the side effects that we see here on our clients every day, the government is even fearful of taking responsibility for these effects, why are they uncomfortable doing so if they are sure of what they are telling us to jab in, ……by the way do you also realize that AstraZeneca has been branded dangerous in Norway and other developed countries (male informant, unvaccinated)*.

Health workers were also concerned about the efficacy and effectiveness of the vaccine because it was reportedly manufactured in India, a country that had registered a high Covid-19 prevalence and resurgence waves in spite of the vaccination with AstraZeneca. Additionally, from their own experience, these health workers had reportedly seen many previously vaccinated colleagues turn up with Covid, some reportedly died from severe Covid-19 while Covid-19 patients with no history of Covid-19 vaccination had less severe Covid-19 illness.

*“…… India where this vaccine is being manufactured still has a high Covid-19 prevalence and has also had several resurgences in spite of people being vaccinated………… …….even here in the hospital our colleagues who got the Covid-19 jab sometimes turn up or even die of Covid-19 while some of us who never vaccinated have never gotten or only get mild symptoms which raises the question of whether this vaccine prevents or accelerates severe Covid-19” (male informant unvaccinated)*.

### Fear of Real or perceived side effects

Fear of side effects the health workers had seen among clients who turned up after vaccination was a barrier for them taking up the vaccine. A few respondents reported they had received several patients come back days or weeks post vaccination with high fevers, swollen arms, severe headaches and difficulty in breathing. Other side effects which although perceived or read from social media scared health workers from getting vaccinated since they neither believed nor dismissed them given the less scientific evidence they had. The perceived effects included the vaccine causing infertility, impotence, body magnetism, vaginal bleeding and brown hair.

*“…*.. *we have seen many patients come back here post vaccination with severe illness, difficulty in breathing or even death…*..*this has not happened commonly with other vaccines we normally get so as a matter of fact, some of us think we are better off not vaccinating than taking the risk of a jab” (female informant, un vaccinated)*.

*“There is also a lot of information especially about this vaccine causing infertility, impotence, magnetism, severe bleeding in women or even hair turning brown………unfortunately, we don’t even have scientific evidence to the contrary so we don’t know what to believe really*.*” (female informant, unvaccinated)*.

### Insufficient sensitization of the health workers amidst vaccine negative publicity

A few health workers were hesitant to take up the vaccine because they lacked sufficient knowledge about it. The government had not reportedly done enough sensitization to the health workers especially on the components, nature, development process, side effects and/or benefits of taking up the vaccine. There were no clear eligibility criteria, indication or contra indications to the vaccine. They were not also reportedly sensitized enough on the other vaccine alternatives and why government was only bent on AstraZeneca. This was against the background that there was a lot of negative publicity about the Covid-19 vaccine including but not limited to misinformation and distortion of scientific facts on the internet and other social media platforms about how dangerous the vaccine was hence casting more doubt about the vaccine.

*“Information about this vaccine is so scanty, there has been no proper sensitization of the workers but just government ordering and threatening us with losing jobs……*.*what is in this vaccine,… what are the benefits of getting it, what is the eligibility criteria…*., *what are the alternatives, how about all the allegations we here about, when will government come out to clarify these issues. We can’t just ignore these pockets of information so…caution”. (male informant, unvaccinated)*.

### Risk denial

Lack of Covid-19 related fear or death was frequently reported by many respondents as one of the reasons some health workers did not take up the vaccine. Several workers believed in their own natural immunity given they had been on the Covid-19 wards, treated many Covid-19 patients but only got mild symptoms of the disease or none at all. Some of them actually retorted that strict observance to standard operating procedures was much safer and less hazardous than taking the up the vaccine.

*“I have been here during the first and second wave, treated so many Covid-19 patients…. but for us some just get mild symptoms or other never at all. So we trust in our natural immunity……*..*besides, some of us believe that strictly observing the standard procedures is much safer than taking the vaccine” (female informant, unvaccinated)*.

### System inhibition factors

A few respondents reported that there were many system gaps in the processes, functions and tools designed to get the health workers accept the Covid-19 vaccine. The gaps included government insistence that all workers take up the vaccine without due diligence which they perceived as coercion in their view, vaccinating them without prior testing to see if they had not been exposed in the first place, few vaccination posts making it expensive to access, putting vaccination centres within the very hospitals that had Covid-19 patients and hence a risk of exposure for those seeking vaccination among others.

*“Why does the government insist that we all get vaccinated more over without even testing us to see if we have had Covid-19 already…*..*you see people lining up in crowds to get the vaccine, moreover the same hospital with Covid-19 patients, isn’t this a super spreader really”. (female informant, vaccinated)*.

### Religious beliefs against vaccination

Some health workers intimated that their religious beliefs did not agree with the notion of vaccines as prevention. Such faiths defined prevention of illness as a preserve of God and not vaccines. They therefore saw no need for the vaccine other than may be doing it to please their superiors or even secure their jobs.

*“Our religion believes it’s only the will of God that keeps us alive. Those vaccines are just a waste of time, I have many friends in this hospital who just pray every morning not to get Covid-19 before they go to the Covid-19 ward and believe me for the two waves they have never got it …as for me, I just got it (the vaccine) to secure my job because we are told that soon anyone who is not vaccinated will not enter that hospital gate. Its prayers, prayers and the Lord listens” (male informant, vaccinated)*.

## 4.0 Discussion

Our findings indicate that the health workers vaccine hesitancy was a function of multiple ecological/cross cutting themes including; gross lack of trust in the vaccine, fear of real/ perceived side effects and insufficient sensitization of health workers amidst vaccine negative publicity. The other cross cutting themes were risk denial, system inhibition factors and religious beliefs against vaccination.

Health workers lacked trust in the Covid-19 vaccine especially in regard to its production processes, the types, need for consent, contents, efficacy, effectiveness, contra-indications and long term safety. They also feared real or perceived side effects of the vaccine based on what they saw and interpreted in their practice and the confusing negative publicity or misinformation over the internet and social media platforms which they neither believed nor dispelled because of lack of relevant scientific knowledge about the Covid-19 vaccine. All these tenets are in real sense functions of misinformation or lack of sufficient knowledge/information by the health workers about the Covid-19 vaccine. Acceptance of a medical product, service or intervention is dependent on how much the recipients know, understand and trust it in terms of safety, benefit or risk reduction among others [22-24]. Knowledge is power and a foundation for appropriate decision making, especially in making health choices. Governments involvement and or sensitization of the health workers in understanding the vaccine trial protocols, program design and the benefits versus risks for them taking up the vaccine was therefore crucial to their decision making [25, 26]. The health workers saw more risks in taking up the vaccine than the benefits accrued to it. Lack of adequate knowledge, involvement and or sensitization of beneficiaries of health interventions as barriers to vaccine acceptance has been established in other studies in Europe and Africa [27-29].

The concept of trust as a pillar in successful health interventions including the Covid-19 vaccine has been amplified by the health belief model, the WHO and other studies on barriers to vaccine acceptance in high and low income settings [6, 12, 30, 31]. Fear of side effects as a barrier to uptake of vaccines has also been established by studies in Europe, Asia and Africa [32-35].

Health workers believing in their own natural immunity and/or just adhering to the standard operating procedures as protective enough against the Covid-19 virus is indicative of lack of risk susceptibility. In actual sense the health workers did not perceive themselves as being at risk of contracting Covid-19 at all. The risk denial may in part be attributed to the real time evidence the health workers saw on the ground between themselves (unvaccinated but healthy) and their colleagues who had been vaccinated but still got Covid-19. The concept of risk denial or risk perception has been found as a barrier to safer health behavioral change because individuals in denial find confidence in scapegoating, self-confidence, false or perceived confidence and comparison between risks [36]. This finding is similar to other studies about Covid-19 or other ailments that require behavioral/attitudinal change where risk denial was established as a barrier to acceptance of an intervention for positive change [37-40].

Government does not seem to have properly explained to the health workers why it was important that every health worker had to get vaccinated immediately even if it meant without testing because of the pandemic status and available resources at that moment in time. As well, the health workers did not seem to appreciate why there were few vaccination posts at that time and why the vaccination was being conducted in the hospital premises that had Covid-19 patients which in their view exposed them to the Covid-19 virus. While these scenarios could probably have been due to the urgency of starting up the vaccination program despite the few vaccines and other resources available, lack of affordable testing kits and refrigeration settings only at the hospitals, the targeted recipients (health workers) lacked a proper awareness/understanding of this appropriate delivery strategy by government at that time hence the hesitancy. Raising knowledge and awareness on health program strategies to recipients is vital to acceptance of the program [41]. Awareness and knowledge about vaccine interventions and their objectives and processes as drivers to acceptance has been established in other studies [42, 43]

Religious beliefs were reported as barriers to the Covid-19 vaccine uptake. Like many other health interventions, there has sometimes been an ideological clash between science and divine healing [44, 45]. Such individuals have been primed by their faith that calamities or pandemics are dawned on us by God and can only be removed by the will of God only. Although sometimes this happens in minority religious groups, they often have a negative multiplier prohibitive effect even to other members of the societies they live in [46]. Evidence of religious beliefs as a barrier to uptake of vaccines has been similarly established in other studies in the United States, Europe, Asia and Africa [47-50]

## 5.0 Conclusions/Recommendations

The government should consider health workers context specific tools/guides for dissemination of information, education and communication about the Covid-19 vaccine. The tools should address awareness on vaccine trial processes and why there was need for emergency authorization of the vaccine, the vaccine types, contents, efficacy, and effectiveness. The tool should also explain why consent for this vaccine is important, the vaccine benefits versus risks, side effects and their prevention or mitigation. Peers who have taken up the vaccine and survived Covid-19 or got a mild form of the disease could also be used to elicit positive peer influence about the vaccine amongst health workers. This could be done through the health worker’s social media platforms, union or association websites, personal statements, editorials or other media. This strategy is likely to elicit trust in the vaccine, minimize fear of side effects and increase risk susceptibility awareness and hence uptake among the health workers. Increasing awareness, information, education and communication about health interventions has been found to improve their uptake [51, 52]. Positive peer influence has also been found useful in improving service uptake in many other interventions [53].

The government could also consider conducting sustained media campaigns either in the mainstream or social media to not only explain the safety and benefits of one taking up a Covid-19 vaccine, but also dispel the misinformation about the vaccine. Using people who got the vaccine and survived Covid-19, religious leaders and other opinion leaders in these campaigns could bolster the effort especially among the faith based vaccine hesitant health workers.

To address system inhibition factors, government could consider context specific dialogue series with health workers. This could be done at the different health system levels from the village health teams, to the national referral hospitals, health workers unions’ or regulatory bodies. The dialogue should aim at helping health workers understand the rationale for insisting that all of them get the vaccine jab and not merely coercion, why they were initially being vaccinated without prior Covid-19 testing ie lack of affordable testing kits and the reasons for the few vaccination centres. The government should ensure that other vaccine types in addition to AstraZeneca are accessible and available at every level of health care. If possible, vaccination posts could also be increased and set outside hospital settings such as schools or play grounds to reduce the risk of exposure. Dialogue with recipients of a service has been found to increase acceptance in many health and non-health interventions [54, 55].

### Study limitations

As it is with qualitative studies, our deductions are purely based on narratives from the respondents with no statistical inferences. The fact that the interviewees themselves lived this life and volunteered the information, however, strengthens the concept that the data derived from the interviews was objective to the topic of the study. Additionally, we also triangulated our data collection methods (FGDs, IDIs) and conducted our analysis iteratively. This helped us to check for consistency and contradictions inside and across the groups and interviewees. Also, our team was multi-disciplinary, grounded in the collection and analysis and had a good contextual understanding of aspects relating to uptake of the Covid-19 vaccine in the study settings. We therefore feel that the content analysis employed for this study has achieved appropriate in-depth analysis for the purpose of the study.

## Data Availability

All data produced in the present study are available upon reasonable request to the authors

